# Evaluating the metabolic effects of neoadjuvant treatment in clear cell renal cell carcinoma using hyperpolarised [1-^13^C]pyruvate MRI

**DOI:** 10.1101/2025.05.22.25328092

**Authors:** Ines Horvat-Menih, Mary A McLean, Jonathan Birchall, Maria Jesus Zamora Morales, Marta Wylot, Stephan Ursprung, Ramona Woitek, Eva Serrao, Ashley Grimmer, Elizabeth Latimer, Alixander S Khan, Andrew N Priest, Andrew B Gill, Joshua D Kaggie, Martin J Graves, Tristan Barrett, James MS Wason, Helen Mossop, Martin Thomas, Sulekha Said, Anne Y Warren, Kate Fife, Tim Eisen, Athena Matakidou, Will Ince, Brent O’Carrigan, James Jones, Sarah J Welsh, Thomas J Mitchell, James N Armitage, Antony CP Riddick, Grant D Stewart, Ferdia A Gallagher

## Abstract

Despite recent advances, ∼50% of people developing renal cell carcinoma (RCC) will die of the disease. The development of new neoadjuvant therapeutic strategies requires reliable companion biomarkers to measure early and successful response to treatment. Tumour size changes are often late markers of response, but novel imaging-based biomarkers may be more accurate for treatment response prediction.

Here we evaluated the potential of hyperpolarised carbon-13 MRI (HP ^13^C-MRI) as an emerging clinical imaging technique for assessing response to neoadjuvant treatment in RCC, as part of the WIndow of opportunity in REnal cancer (WIRE) trial.

The change in LAC/PYR ratio following treatment was variable across the four patients (mean±S.D. %change = +6±27%). LAC/PYR decreased in the patient treated with cediranib monotherapy (−21%), and in one of the patients receiving combination treatment (−14%). A higher LAC/PYR ratio post-treatment was observed in the second patient receiving combination treatment (+21%) and in the patient receiving olaparib monotherapy (+35%).

This is the first study to evaluate the potential of clinical HP ^13^C-MRI in assessing early treatment response in renal cancer, which detected metabolic changes following treatment in the absence of significant changes in tumour size. Future studies should assess this finding in larger patient cohorts.

**Patient summary:** In this study we used an emerging clinical imaging technique, called hyperpolarised carbon-13 MRI, to visualise how kidney cancer changes with drug treatment before surgery. The method visualised rapid changes in cancer metabolism before substantial changes were seen in tumour size, the latter being the conventional method for detecting response to treatment. Hyperpolarised carbon-13 MRI holds promise in informing clinicians which cancers have successfully responded, and which may benefit from a change in treatment.

## Introduction

Renal cell carcinoma (RCC) is the most common kidney malignancy with an increasing incidence globally, but with no decline in mortality rates^1^. Surgery with curative intent is the treatment of choice for clinically-fit patients whose disease burden is amenable to resection, yet ∼30% experience recurrence^1,2^. Neoadjuvant approaches may allow more rapid treatment of patients with aggressive localised disease whilst potentially minimising the need for adjuvant therapy after surgery^3^.

A particular challenge for studies evaluating neoadjuvant therapy is the lack of validated predictive biomarkers in RCC, which delays the advancement of novel agents^3^. Currently the treatment response is determined using the RECIST 1.1 criteria (Response Evaluation Criteria in Solid Tumours), which are based on the change in size of target lesions^4^. However, not only is evidence lacking for the use of RECIST-defined progression as a clinically valid endpoint for treatment modification, but also morphologic changes alone may be insufficient for capturing response to novel targeted therapies^4^.

Hyperpolarised [1-^13^C]pyruvate MRI (HP ^13^C-MRI) is a novel clinical imaging technique to probe metabolic conversion of injected hyperpolarised [1-^13^C]pyruvate tracer to [1-^13^C]lactate. A decrease in tumour lactate labelling has been demonstrated after several weeks of treatment in prostate cancer^5^, and furthermore, the technique detected response to neoadjuvant therapy in breast cancer after only 7-11 days, which outperformed conventional MRI in distinguishing pathological complete response from non-responders^6^. In renal cancer, the HP ^13^C-MRI has shown potential for assessing tumour aggressiveness, but has not yet been probed as a marker of treatment response^7^.

WIndow-of-opportunity in REnal cell cancer (WIRE) is a phase II, multi-arm, non-randomised trial which probes the biological mechanism of novel targeted therapies during the interval between diagnosis and surgery (NCT03741426). Here we report the application of HP ^13^C-MRI to probe metabolic treatment response to: cediranib (a tyrosine-kinase inhibitor), olaparib (a poly ADP-ribose polymerase or PARP inhibitor), and both agents in combination. Findings were compared to changes in tumour size, perfusion, and diffusion.

## Results

Details on Patients and Methods are to be found in Supplementary. Four patients with a diagnosis of resectable clear cell RCC (ccRCC) who were recruited to the WIRE trial underwent successful HP ^13^C-MRI before and after treatment. An overview of the clinical characteristics is presented in Suppl. Table S1.

Representative maps of ^13^C-LAC/PYR ratio overlaid on anatomical T_1_w images before and after treatment are presented in Fig. 1. Quantitative results are presented in Suppl. Table S2. The mean LAC/PYR ratio change for all four patients showed a slight increase of +6% post-treatment but with a significant interpatient variation in the S.D. of 27%. Patients 1 and 2 demonstrated a decrease in the tumour LAC/PYR of -21% (baseline: 0.19; post-treatment: 0.15) and -14% (baseline: 0.21; post-treatment: 0.18), while Patients 3 and 4 showed an increase in the tumour LAC/PYR ratio post-treatment of +21% (baseline: 0.07; post-treatment: 0.09) and +35% (baseline: 0.17; post-treatment: 0.23) respectively.

**Figure 1.**
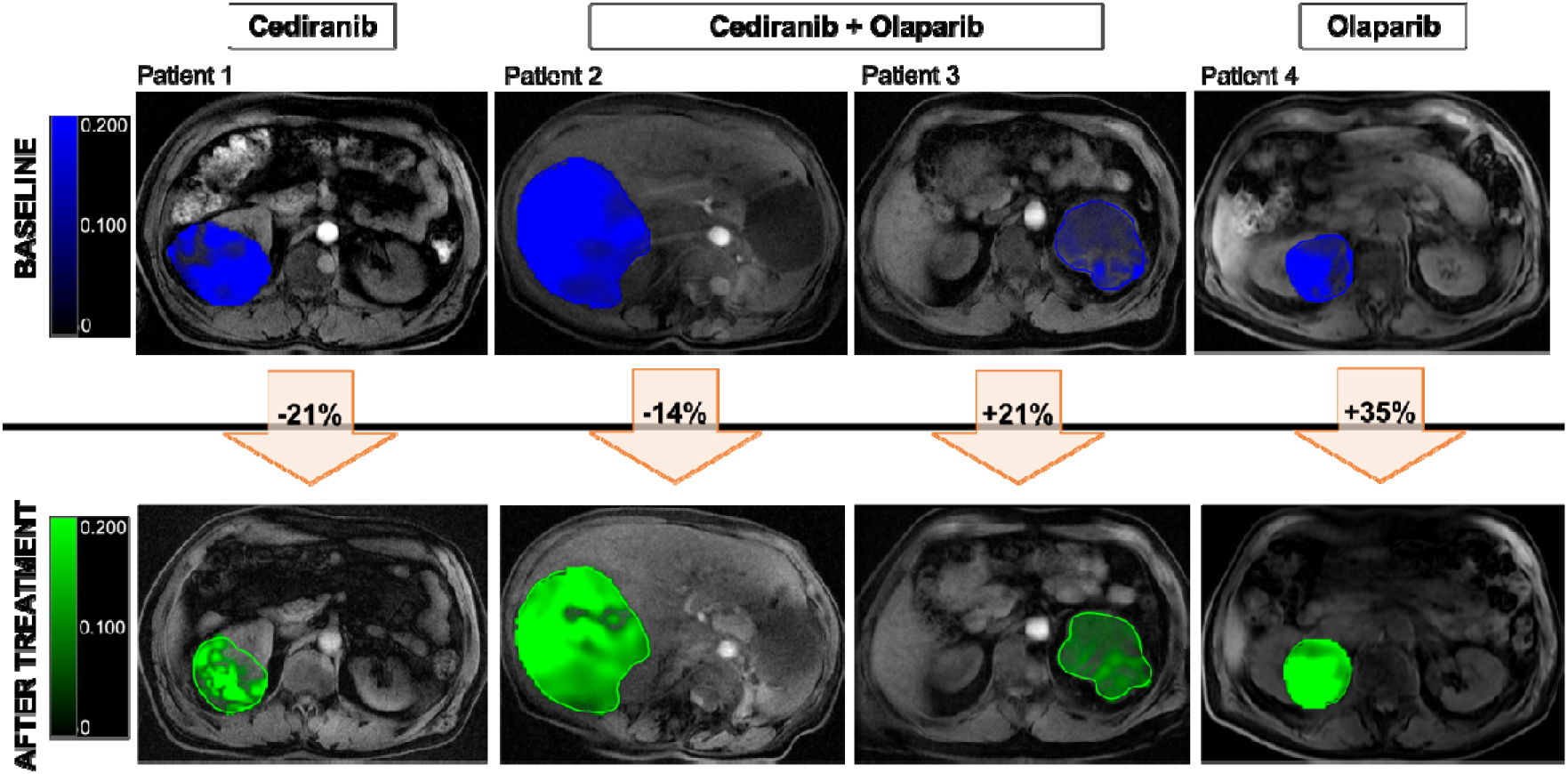
Representative LAC/PYR maps of the four patients overlaid on T_1_w images, with tumour ROIs annotated in blue (baseline) and green (post-treatment). Percentage changes between baseline and post-treatment LAC/PYR ratio presented in arrows for each individual patient. ROI = region of interest.

The perfusion surrogate, *f*_p_, showed the greatest and most consistent changes post-treatment of -31±18%, followed by the hypoxia surrogate, *R*_2_*, of +18±21% (Fig. 2a). The treatment effect on diffusion coefficient, *D*_0_, was variable, where Patients 1 and 3 exhibited a decrease (−14% and -3%, respectively), and Patients 2 and 4 an increase (+4% and +10%, respectively). Only small alterations were observed in tumour diameters, with a mean decrease of -4% from 9.1 cm at baseline to 8.7 cm at post-treatment (S.D. 3%). Mean volumetric changes were greater: -10% over the treatment time course (S.D. 18%); Patient 4 (olaparib only) showed a 13% increase in tumour volume post-treatment. This was associated with greatest increase in *D*_0_ of +10% indicating reduced restriction, the largest decrease in the *f*_p_ of -57%, as well as the highest baseline *R*_2_*, which remained high post-treatment (0% change). Correlative analysis as shown in Fig. 2b, with detailed results in Suppl. Table S3, confirmed a significant relationship between the %changes in tumour volume and *D*_0_ (Pearson r = 0.96, *P* < 0.05), and though non-significant, a strong negative correlation coefficient with *f*_p_ (Pearson r = -0.84, *P* = 0.165), and *R*_2_* (Pearson r = -0.92, *P* = 0.052), respectively. The LAC/PYR changes were independent of the alterations in other imaging metrics.

**Figure 2.**
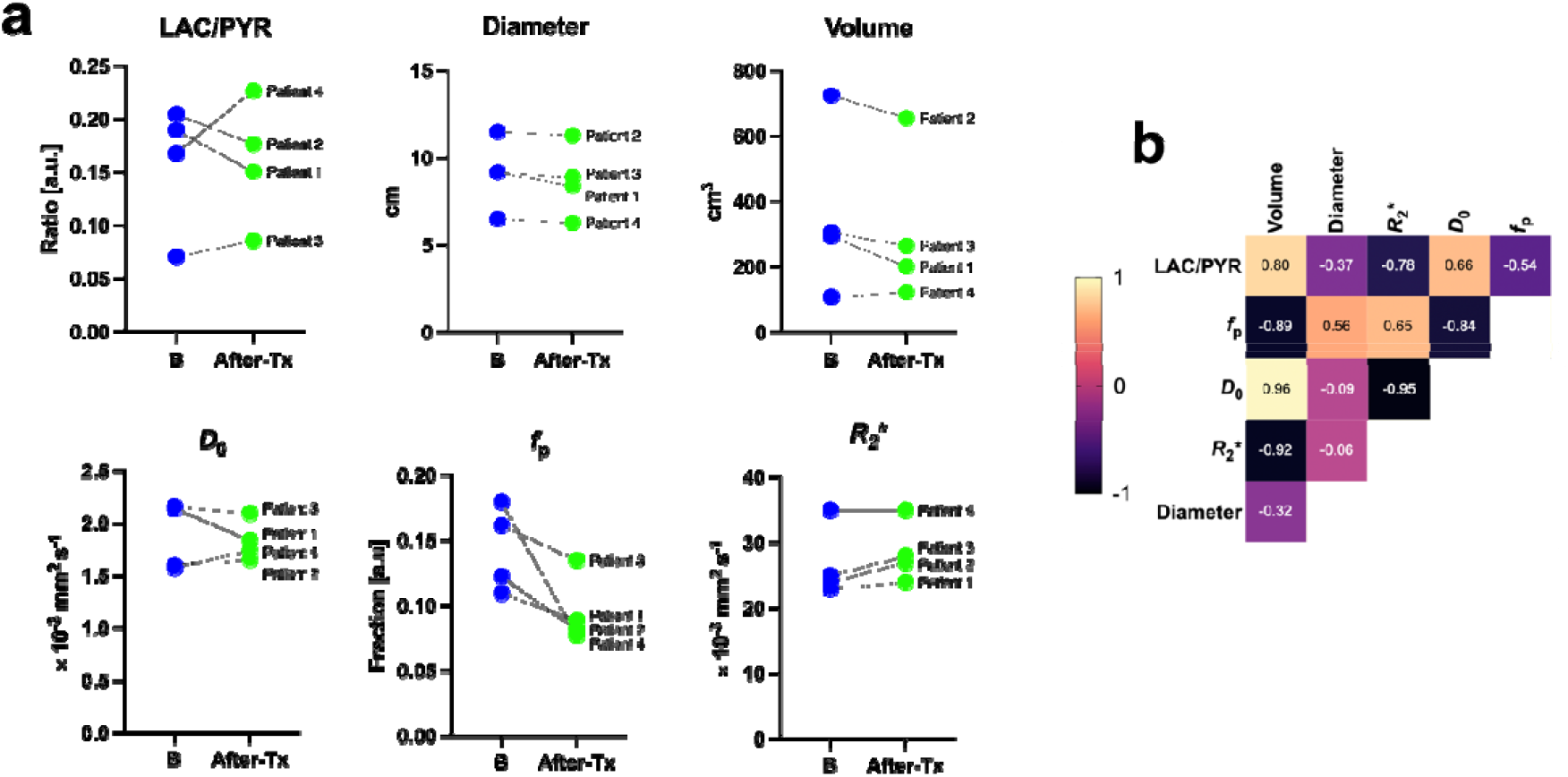
(a) Paired plots representing changes in the imaging parameters pre- (blue) and post-treatment (green). LAC/PYR ratio and *D*_0_ showed variable changes, whereas *f*_p_ and *R*_2_* were most consistently altered post-treatment. (b) Correlation analyses between these imaging parameters presented in a heatmap, labelled with respective Pearson coefficients. B = baseline, After-Tx = after-treatment.

## Discussion

Four patients undergoing neoadjuvant treatment showed variable changes in the tumour LAC/PYR ratio, which occurred in the absence of diameter or volume changes, suggesting that HP ^13^C-MRI may distinguish early metabolic changes before morphological ones can be measured. These results are similar to those reported previously in breast cancer, where an increase in LAC/PYR ratio predicted response to standard-of-care neoadjuvant chemotherapy after 7-11 days^6^.

We have demonstrated previously that the LAC/PYR ratio increased in breast cancer patients undergoing olaparib therapy, irrespective of their response^6^. PARP inhibitors are known to increase intracellular concentrations of nicotinamide adenine dinucleotide (NAD+), an essential co-factor for many redox pathways, which then becomes available as a co-factor for the reaction catalysed by lactate dehydrogenase, promoting the exchange between pyruvate and lactate^8^. An increase in lactate labelling could therefore represent drug engagement with the target^9^. The LAC/PYR ratio decreased in the patient receiving cediranib monotherapy (−21%) and increased in the patient receiving olaparib alone (+35%), with the two patients on combinational therapy showing values between these two values: this supports the suggestion that cediranib and olaparib have opposing effects on tumour metabolism, as well as potential for distinguishing between responders and non-responders in patients receiving olaparib. Furthermore, Patient 4 displayed the greatest decrease in the perfusion fraction (−57%) across all the patients, even though olaparib is not an antiangiogenic agent and this may be related to high tumour hypoxia, as measured by the hypoxia surrogate *R*_2_*, which has been previously suggested to enhance the action of olaparib^10^.

The present study is the first to evaluate the potential of HP ^13^C-MRI in assessing early treatment response in renal cancer. HP ^13^C-MRI detected metabolic changes following treatment in the absence of significant changes in size on clinical proton MRI and larger trials are needed to confirm these findings and to correlate these changes with long-term clinical outcome.

## Data Availability

All data produced in the present study are available upon reasonable request to the authors.

## Acknowledgements

We acknowledge the invaluable feedback by the patient representatives, as well as the administrative and technical support from the Advanced Cancer Imaging and Urological Malignancies programmes, Cancer Research UK (CRUK) Cambridge Centre, and radiographers of the Magnetic Resonance Spectroscopy Unit, Addenbrookes.

