## Supplementary for "Evaluating the metabolic effects of neoadjuvant treatment in clear cell renal cell carcinoma using hyperpolarised [1-^13^C]pyruvate MRI"

***Patients & methods****:*

*Ethics and recruitment*

Patients with renal masses presenting to the Uro-oncology clinic at Addenbrooke’s Hospital, Cambridge University Hospitals NHS Foundation Trust, Cambridge UK, between December 2020 and March 2024, were prospectively recruited and provided written informed consent for the ethically approved clinical trial WIRE (ClinicalTrials.gov: NCT03741426). Inclusion criteria for this imaging report were as follows: age ≥18 years old, clinical suspicion of a renal mass, Eastern Cooperative Oncology Group (ECOG) performance status ≤1, biopsy-proven and surgically resectable ccRCC, successful HP ^13^C-MRI acquisition on pre- and post-treatment imaging. Exclusion criteria were as follows: unsuitability for MRI, significant comorbidities, pregnancy, immunosuppression, previous exposure to tyrosine-kinase and PARP inhibitors. Primary tumour response was assessed according to RECIST 1.1 criteria, comparing baseline and post-treatment imaging before surgery, as per WIRE trial protocol^1^. Tumour stage and grade were evaluated post-surgery, based on TNM (Tumour, Node, Metastasis) classification and WHO/ISUP (World Health Organisation/International Society for Urologic Pathologists) grading system.

*HP ^13^C-MRI*

The ^13^C-pyruvate injection was prepared as described previously^2,3^. HP ^13^C-MR images were acquired on a 3 T MR system (MR750 or Premier, GE Healthcare, Waukesha WI, USA). On the MR750, ^13^C-tuned clamshell transmit and 8-channel array receive coils were used (Rapid Biomedical, Rimpar, Germany) and on the Premier, a 4-rung birdcage ^13^C transmit/receive coil was used (Rapid Biomedical, Rimpar, Germany) with an additional 16-channel flexible AIR coil array for reception (Neocoil, Pewaukee WI, USA). At 12 s after the start of injection, a spiral imaging sequence was used to acquire 5 slices, each 3 cm thick with 3 mm gaps and with the following parameters: 340x340 mm^2^ field of view (FOV) with matrix size 40x40, temporal resolution 4s, 20 timepoints (repetition time 1 s), reconstructed in-plane resolution 128x128 and true voxel resolution 17x17x30 mm^3^. Individual metabolites were imaged following spectral-spatial excitation^4^ in a 4-step cycle following the scheme: pyruvate 10°, lactate 40°, pyruvate 10°, alanine 40°.

Acquisition and reconstruction of HP ^13^C-MRI data was performed on the scanner using the Multinuclear Spectroscopy Research Package (GE Healthcare, Waukesha WI, USA). Metabolite images from individual timepoints were summed together offline using an in-house MATLAB (MathWorks Inc., Natick MA, USA) script, and the complex imaging data from the 8-channels of the abdominal coil were combined using a singular value decomposition approach. Masking was performed to only include voxels where the SNR from the summed area under the curve (AUC) of ^13^C-lactate+pyruvate was ≥5. DICOM format metabolite images were written for each slice, including total ^13^C signal-to-noise ratio (SNR) summing over all metabolites, summed ^13^C-pyruvate, summed ^13^C-lactate, and normalised ^13^C-lactate/pyruvate ratio (LAC/PYR) i.e.^13^C-lactate/(^13^C-lactate+pyruvate).

Analysis of metabolite maps was performed in OsiriX Lite v.12.0.3 (Pixmeo SARL, Switzerland). Tumour regions of interest (ROIs) were drawn on axial T_1_w LAVA-Flex images and were then propagated to the metabolic maps.

*Proton MRI*

Proton MRI (^1^H-MRI) was acquired immediately after HP ^13^C-MRI by replacing the carbon-tuned coils with the 32-channel cardiac array ^1^H coil (GE Healthcare, Waukesha WI, USA) and imaging the study participant in the same 3 T MRI scanner. The protocol included T_1_w, T_2_w, *R*_2_* mapping, and IVIM (intravoxel incoherent motion) diffusion-weighted imaging (DWI) sequences as described previously^3^. Images were processed using GE and in-house developed MATLAB-implemented software to extract the biomarker maps. Specifically, IVIM-DWI data was motion corrected across all b-values, and maps of diffusion coefficient, *D*_0_, were computed based on nonlinear fits to multi-value diffusion images.

For quantification, the acquired maps underwent resampling and manual registration to the axial T_1_w LAVA-Flex images using ITK-SNAP 3.8 software (University of Pennsylvania, USA). Tumour ROIs were drawn in OsiriX Lite v.12.0.3 (Pixmeo SARL, Switzerland) on coronal T_1_w LAVA-Flex and propagated to manually co-registered maps. Quantitative parameters were calculated from voxel intensities and reported as mean±standard deviation (S.D.).

*Statistical analysis*

Statistical analysis was performed in GraphPad Prism v.10.0.2 (Dotmatics, Boston MA, USA). Mean and S.D. summarised the normally distributed data, as probed with a Shapiro-Wilk test for normality. The Pearson correlation coefficient was calculated to evaluate for dependency between imaging parameters. *P* < 0.05 was deemed as a cut-off for statistical significance.

**Data availability:** Data are available for bona fide researchers who request it from the authors.

**Code availability**: Custom computer code was used to generate the results for imaging, and is available upon request.

Supplementary Table S1: Patient characteristics.

|  | Age at detection [years]; Sex | Tumour diameter at detection | Presentation at detection (relevant comorbidities); ECOG status | Treatment (days) | Preoperative RECIST 1.1 | Stage and grade of the tumour |
| --- | --- | --- | --- | --- | --- | --- |
| **Patient 1** | In their 50’s, M | 9.2 cm | Right flank pain (hypertension);  ECOG 0 | Cediranib, 25d | Stable disease | ypT3a pN0 M0;  Grade 4 |
| **Patient 2** | In their 60’s, F | 11.5 cm | Haematuria (cachexia, anaemia);  ECOG 1 | Cediranib + olaparib, 26d | Stable disease | ypT3b N0 M1 (lungs);  Grade 4 |
| **Patient 3** | In their 50’s, M | 9.2 cm | Incidental (hypertension);  ECOG 0 | Cediranib + olaparib, 22d | Stable disease | ypT3a pN0 M0;  Grade 3 |
| **Patient 4** | In their 70’s, M | 6.5 cm | Weight loss (hypertension, pulmonary embolism);  ECOG 1 | Olaparib, 28d | Stable disease | ypT3b pN0 M0;  Grade 3 |

Supplementary Table S2: Quantitative results for imaging parameters extracted from tumour regions within the four patients.

|  | **Total carbon**  **SNR [a.u]** | **LAC/PYR [a.u.]** | ***f*_p_ [a.u.]** | ***R*_2_* [s^-1^]** | **D0**  **[×10^-3^ mm^2^ s^-1^]** | **Diameter [cm]** | **Volume [cm^3^]** |
| --- | --- | --- | --- | --- | --- | --- | --- |
| **Patient 1, baseline** | 18 | 0.19 | 0.110 | 23 | 2.15 | 9.2 | 293 |
| **Patient 1, post-treatment** | 19 | 0.15 | 0.089 | 34 | 1.84 | 8.4 | 201 |
| **Patient 2, baseline** | 30 | 0.21 | 0.123 | 24 | 1.60 | 11.5 | 726 |
| **Patient 2, post-treatment** | 20 | 0.18 | 0.082 | 27 | 1.66 | 11.3 | 656 |
| **Patient 3, baseline** | 29 | 0.07 | 0.162 | 25 | 2.16 | 9.2 | 306 |
| **Patient 3, post-treatment** | 23 | 0.09 | 0.135 | 28 | 2.10 | 8.9 | 266 |
| **Patient 4, baseline** | 14 | 0.17 | 0.180 | 35 | 1.58 | 6.5 | 109 |
| **Patient 4, post-treatment** | 24 | 0.23 | 0.078 | 35 | 1.74 | 6.3 | 124 |
| **Mean, baseline** | 23 | 0.16 | 0.144 | 27 | 1.87 | 9.1 | 359 |
| **Mean, post-treatment** | 22 | 0.16 | 0.096 | 31 | 1.84 | 8.7 | 312 |
| **S.D., baseline** | 8 | 0.06 | 0.033 | 6 | 0.33 | 2.0 | 261 |
| **S.D., post-treatment** | 2 | 0.06 | 0.026 | 4 | 0.19 | 2.1 | 237 |
| **%Changes of imaging parameters** | | | | | | | |
|  | **Patient 1** | -21 | -19 | 48 | -14 | -9 | -31 |
|  | **Patient 2** | -14 | -33 | 13 | 4 | -2 | -10 |
|  | **Patient 3** | 21 | -17 | 12 | -3 | -3 | -13 |
|  | **Patient 4** | 35 | -57 | 0 | 10 | -3 | 13 |
|  | **Mean %changes** | 6 | -31 | 18 | -1 | -4 | -10 |
|  | **S.D. %change** | 27 | 18 | 21 | 10 | 3 | 18 |

Supplementary Table S3: Correlative analysis of %change of imaging parameters extracted from tumour regions within the four patients.

| **Pearson r** | **Volume** | **Diameter** | ***R*_2_*** | ***D*_0_** | ***f*_p_** |
| --- | --- | --- | --- | --- | --- |
| **LAC/PYR** | 0.80 | 0.47 | -0.78 | 0.66 | -0.54 |
| ***f*_p_** | -0.89 | -0.44 | 0.65 | -0.84 |  |
| ***D*_0_** | 0.96 | 0.86 | -0.95 |  |  |
| ***R*_2_*** | -0.92 | -0.91 |  |  |  |
| **Diameter** | 0.71 |  |  |  |  |
| ***P* values** | **Volume** | **Diameter** | ***R*_2_*** | ***D*_0_** | ***f*_p_** |
| **LAC/PYR** | 0.196 | 0.525 | 0.221 | 0.337 | 0.458 |
| ***f*_p_** | 0.111 | 0.560 | 0.346 | 0.165 |  |
| ***D*_0_** | 0.039 | 0.145 | 0.052 |  |  |
| ***R*_2_*** | 0.084 | 0.090 |  |  |  |
| **Diameter** | 0.287 |  |  |  |  |

1.	Ursprung S, Mossop H, Gallagher FA, et al. The WIRE study a phase II, multi-arm, multi-centre, non-randomised window-of-opportunity clinical trial platform using a Bayesian adaptive design for proof-of-mechanism of novel treatment strategies in operable renal cell cancer – a study protocol. *BMC Cancer*. 2021;21(1):1238. doi:10.1186/s12885-021-08965-4

2.	Gallagher FA, Woitek R, McLean MA, et al. Imaging breast cancer using hyperpolarized carbon-13 MRI. *Proc Natl Acad Sci USA*. 2020;117(4):2092-2098. doi:10.1073/pnas.1913841117

3.	Ursprung S, Woitek R, McLean MA, et al. Hyperpolarized 13C-Pyruvate Metabolism as a Surrogate for Tumor Grade and Poor Outcome in Renal Cell Carcinoma—A Proof of Principle Study. *Cancers*. 2022;14(2):335. doi:10.3390/cancers14020335

4.	Schulte RF, Sperl JI, Weidl E, et al. Saturation‐recovery metabolic‐exchange rate imaging with hyperpolarized [1‐ ^13^ C] pyruvate using spectral‐spatial excitation. *Magnetic Resonance in Med*. 2013;69(5):1209-1216. doi:10.1002/mrm.24353
